# Radiomics-Based Lung Nodule Classification with Stochastic Search Variable Selection and Bayesian Logistic Regression

**DOI:** 10.1101/2025.04.29.25326704

**Authors:** Isaac Lam

## Abstract

**Background:** Radiomics is an emerging field in medical science that involves extracting quantitative features from medical images using data characterization algorithms. These features, known as radiomic features, provide a quantitative approach to medical image analysis. In the context of lung cancer, radiomics-based approaches are transforming disease management by improving early detection, diagnosis, prognosis, and treatment decision-making.

**Objective:** This study aimed to explore the utility of Bayesian methods, specifically Stochastic Variable Selection and Shrinkage (SVSS) and Bayesian logistic regression, in the radiomics-based classification of small lung nodules with limited training data.

**Methods:** The Bayesian approach was compared to frequentist Lasso logistic regression on the test set. The performance of both methods was evaluated to determine their viability in the classification of small lung nodules.

**Results:** The Bayesian approach matched the performance of frequentist Lasso logistic regression on the test set, demonstrating its viability as an alternative approach. Annulus GLCM Entrop LLL was consistently identified as a feature positively influencing small lung nodule malignancy prediction across multiple models. This finding enhances confidence in the effect of this feature, suggesting that future Bayesian analyses can incorporate this information for greater reliability in feature selection and coefficient estimates.

**Conclusion:** This study highlights the potential of Bayesian methods to address the challenges of limited data in medical image analysis, offering a robust alternative to traditional statistical approaches and contributing to improved clinical decision-making.

## Introduction

Radiomics is an emerging field in medical science that involves extracting a large number of quantitative features from medical images using data characterization algorithms. These features, known as radiomic features, contain textural information such as spatial distribution of signal intensities and pixel interrelationships that can provide a quantitative approach to medical image analysis [8].

In lung cancer research, radiomics-based methods are revolutionizing disease management by enhancing early detection, diagnosis, prognosis, and treatment decision-making. A critical clinical metric for evaluating cancer risk is the presence of lung nodules, with the size of these nodules being a significant factor [4]. The diameter of a nodule is measured to categorize it as either small or large.

The radiomics-based approach has emerged as a cutting-edge method for enhancing lung cancer risk prediction. Extracting radiomic features from medical images and analyzing them with machine learning (ML) models offers a non-invasive non-invasive complement to CT scans for classifying of lung nodules. This approach has important implications for clinical decision making, as it can enhance diagnostic and predictive accuracy, particularly for indeterminate nodules [6]. Given that radiomic features can be represented in high-dimensional datasets containing hundreds or thousands of features, previous research have employed feature selection methods such as Lasso regression [2], random forest feature ranking [6], and principal component analysis for dimensionality reduction [3]. Others have selected specific features based on the morphological characteristics they represent [5]. However, to my best knowledge, stochastic search variable selection (SSVS) has yet to be implemented in this context.

Medical image analysis often faces the challenge of limited data availability. This scarcity can be attributed to the rarity of certain medical conditions and the high cost of imaging procedures. Traditional statistical methods struggle with small sample sizes, leading to unstable and unreliable estimates. In contrast, Bayesian methods, such as Bayesian logistic regression and Stochastic Variable Selection and Shrinkage (SVSS), offer a robust solution by incorporating prior information into the model. This integration stabilizes estimates and enhances model performance, making Bayesian approaches particularly valuable in medical image classification tasks [7]. Additionally, these Bayesian methods improve interpretability by providing a range of probability estimates for each parameter, which offer a more nuanced understanding of the model’s predictions.

The objective of this study is to integrate SSVS and bayesian logistic regression into radiomics-based classification of small lung nodules. By leveraging the strengths of Bayesian methods, this study aims to enhance the accuracy and reliability of lung nodule classification, ultimately contributing to improved clinical decision-making and patient outcomes.

### Data Source

The dataset utilized in this study is publicly accessible and was initially collected and annotated by Hunter et al. [2]. The original dataset consists of CT images of lung nodules, which were processed to extract radiomic features using TexLab2.0. The final dataset comprises 736 images, each containing 1998 radiomic features that quantitatively describe the shape, texture, and intensity of the nodules. More details regarding the data collection, annotation procedures, and radiomic feature extraction can be found in the paper by Hunter et al. [2].

## Methodology

The methodology employed in this study is adapted from Hunter et al. [2]. To evaluate the effectiveness of SVSS and bayesian logistic regression on limited data, 50 images were utilized for training while the remaining 686 were set aside for testing. All 1998 radiomic features underwent Z-score normalization, defined as 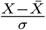.

To mitigate computational demands, an initial layer of feature selection was performed using univariate frequentist logistic regression to identify significant features. Features were deemed significant if they did not exceed the adjusted critical value after false discovery rate (FDR) correction, utilizing the Benjamini-Hochberg procedure with an FDR set to 0.1. This process identified 316 significant features. Subsequently, Stochastic Variational Subset Selection (SVSS) was applied to the remaining dataset to select the final model features. The top three features with the greatest posterior probabilities from SVSS were selected.

Bayesian logistic regression models were developed to predict malignancy using radiomic features selected by SVSS. Four distinct Bayesian logistic regression models were constructed, each differing by the precision parameter *τ* for the normal priors applied to the coefficients and intercept. The specific *τ* values used were 0.01, 0.1, 1, and 10. Pareto-smoothed importance sampling leave-one-out cross-validation (PSIS-LOO-CV) was employed to determine the most suitable model.

The performance of the optimal bayseian logistic regression model was compared against a baseline frequentist Lasso logistic regression model, which was constructed using the significant features identified from the univariate logistic regressions. The regularization hyperparameter *C* for the Lasso model was tuned in a five fold cross-validation loop, optimizing for the area under receiver operating characteristic curve (AUROC).

### Model Evaluation

Receiver Operating Characteristic (ROC) curves were employed to evaluate the performance of both models. AUROC was utilized as the primary metric for assessment. To estimate the 95% confidence intervals for the AUROC, resampling was conducted using 1000 bootstrap iterations. Threshold-based performance metrics, including accuracy, precision, recall, and F1-Scores, were calculated based on the ROC cutoff that maximized the Youden index on the training set. The Youden index, which provides equal weight to both sensitivity and specificity, was selected as a balanced metric for this purpose.

### Stochastic Search Variable Selection (SSVS)

SSVS aims to identify which predictors should be included in the model. It does this by introducing binary indicators for each predictor, which determine whether a predictor is included or excluded from the model at each sampling iteration in a Markov Chain Monte Carlo (MCMC) simulation.

To perform variable selection, a binary indicator *δ*_*k*_ was introduced for each radiomic feature *x*_*k*_. The indicator *δ*_*k*_ follows a Bernoulli distribution with a probability of 0.0015:

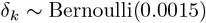

The prior distribution for the coefficients *α*_*k*_ was defined as:

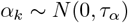

The actual coefficient *β*_*k*_ was then determined by the product of *δ*_*k*_ and *α*_*k*_:

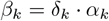

The intercept *β*_0_ was assigned a normal prior distribution:

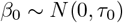

The linear predictor *η*_*i*_ was defined as:

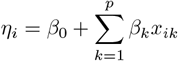

where *p* is the number of predictors, and *i* represents the *i*^th^ data point.

The probability of the binary outcome *y*_*i*_ = 1 was modeled using the sigmoid function:

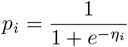

The likelihood of the observed data *y* was specified as a Bernoulli distribution:

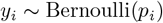

*τ*_*α*_ and *τ*_0_ were set to 1 because smaller values of the precision parameter resulted in divergences during sampling. MCMC methods from the `pymc` python module were used to sample from the posterior distribution of the parameters. The python scripts for model development are available at: https://github.com/isaaclhk/Projects/tree/main/lungnoduleSVSS.

## Results

### Nodule Classification Statistics

Among the 736 samples in the dataset, 377 nodules (51.2%) were classified as malignant, while 356 nodules (48.8%) were identified as benign. In the training set, 26 nodules (52%) were malignant and 24 nodules (48%) were benign. The test set comprised 686 samples, with a similar distribution of 351 malignant nodules (51.2%) and 335 benign nodules (48.8%). For more detailed information on patient and nodule characteristics, refer to the study by Hunter et al. [2].

### Convergence Statistics

A total of 4 chains of 11000 draws were simulated for SVSS, discarding the first 1000 as burn-in. 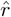 values were 1.0 for all binary indicators *δ*_*k*_. The bulk and tail effective sample size (ESS) for all *δ* exceeded 3000, indicating adequate sampling (Table S1). For each Bayesian logistic regression model, I simulated 4 chains with 6,000 draws each, also discarding the first 1,000 as burn-in. The best performing model had bulk and tail ESS values exceeding 10,000 for all coefficients, with 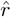 values of 1.0 (see Table S2). Trace plots are provided in Figure S1 in the supplementary materials.

### SVSS

The top three features selected by SVSS and the posterior means of their corresponding indicator parameters *δ*_*k*_ are reported in Table 1.

**Table 1:**
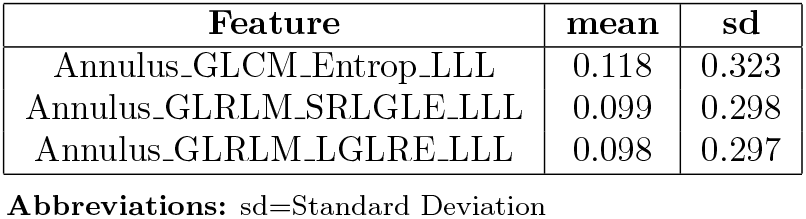
Corresponding posterior mean of the indicator parameter *δ*_*k*_ for each selected feature.

| Feature | mean | sd |
| --- | --- | --- |
| Annulus_GLCM_Entrop_LLL | 0.118 | 0.323 |
| Annulus_GLRLM_SRLGLE_LLL | 0.099 | 0.298 |
| Annulus_GLRLM_LGLRE_LLL | 0.098 | 0.297 |

**Abbreviations:** sd=Standard Deviation

### Bayesian Logistic Regression Model

The model with prior precision parameter *τ* = 1 was identified as the optimal model, achieving the greatest Expected log pointwise predictive density for leave-one-out cross-validation (elpd_loo) value of -26.89. Additionally, this model demonstrated an effective number of parameters (p_loo) of approximately 2 (Table S3).

The posterior distributions of the model coefficients and their highest density intervals (HDIs) are summarized in Table 2. *β*_1_ has a mean of 0.791(95% HDI: -0.324 to 1.859), indicating that it is positively associated with malignancy prediction. In contrast, *β*_2_ and *β*_3_ have means of -0.592(95% HDI: -2.174 to 0.941) and -0.611(95% HDI: -2.165 to 0.951), respectively, suggesting negative influence.

**Table 2:** Coefficients of Bayesian Logistic Regression model with *τ* = 1.

| Coefficient | Mean | SD | HDI 2.5% | HDI 97.5% |
| --- | --- | --- | --- | --- |
| Intercept | -0.107 | 0.342 | -0.772 | 0.557 |
| $\beta_1$ | 0.791 | 0.556 | -0.324 | 1.859 |
| $\beta_2$ | -0.592 | 0.797 | -2.174 | 0.941 |
| $\beta_3$ | -0.611 | 0.799 | -2.165 | 0.951 |

**Abbreviations:** df=Standard Deviation; HDI=Highest Density Interval

### Baseline Lasso Logistic Regression Model

An optimal value for the regularization parameter was determined to be *C* = 0.184. The Lasso method identified 8 features, with their corresponding coefficients detailed in Table 3. Notably, two of the three features selected by SVSS —Annulus_GLRLM_SRLGLE_LLL and Annulus_GLCM_Entropy_LLL— were also chosen by Lasso. However, the third feature, Annulus_GLRLM_LGLRE_LLL, was not selected by Lasso. Similar to Bayesian logistic regression, the Lasso coefficients suggest that Annulus_GLRLM_SRLGLE_LLL has a negative effect on malignancy prediction. Additionally, Annulus_GLCM_Entropy_LLL was predicted by Lasso to increase the risk of malignancy.

**Table 3:**
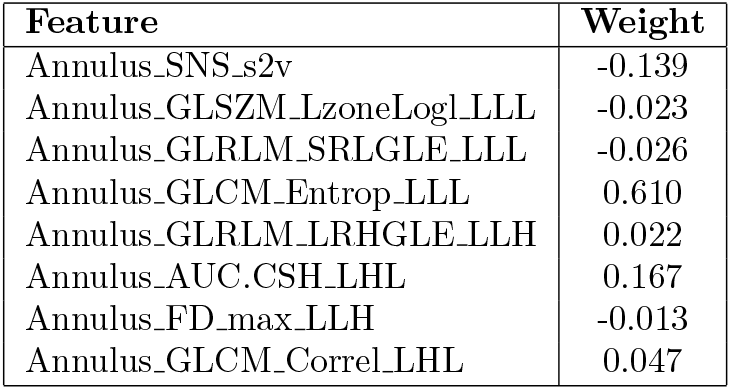
Selected features and their coefficients from Lasso logistic regression.

### Model Evaluation

The selected Bayesian logistic regression model achieved an AUROC score of 0.832 (95% CI: 0.704 to 0.933) on the training set and 0.795 (95% CI: 0.759 to 0.829) on the test set (Figure 1). Additionally, the accuracy score was 0.738 (95% CI: 0.620 to 0.860) on the training set and 0.722 (95% CI: 0.688 to 0.755) on the test set.

**Figure 1.**
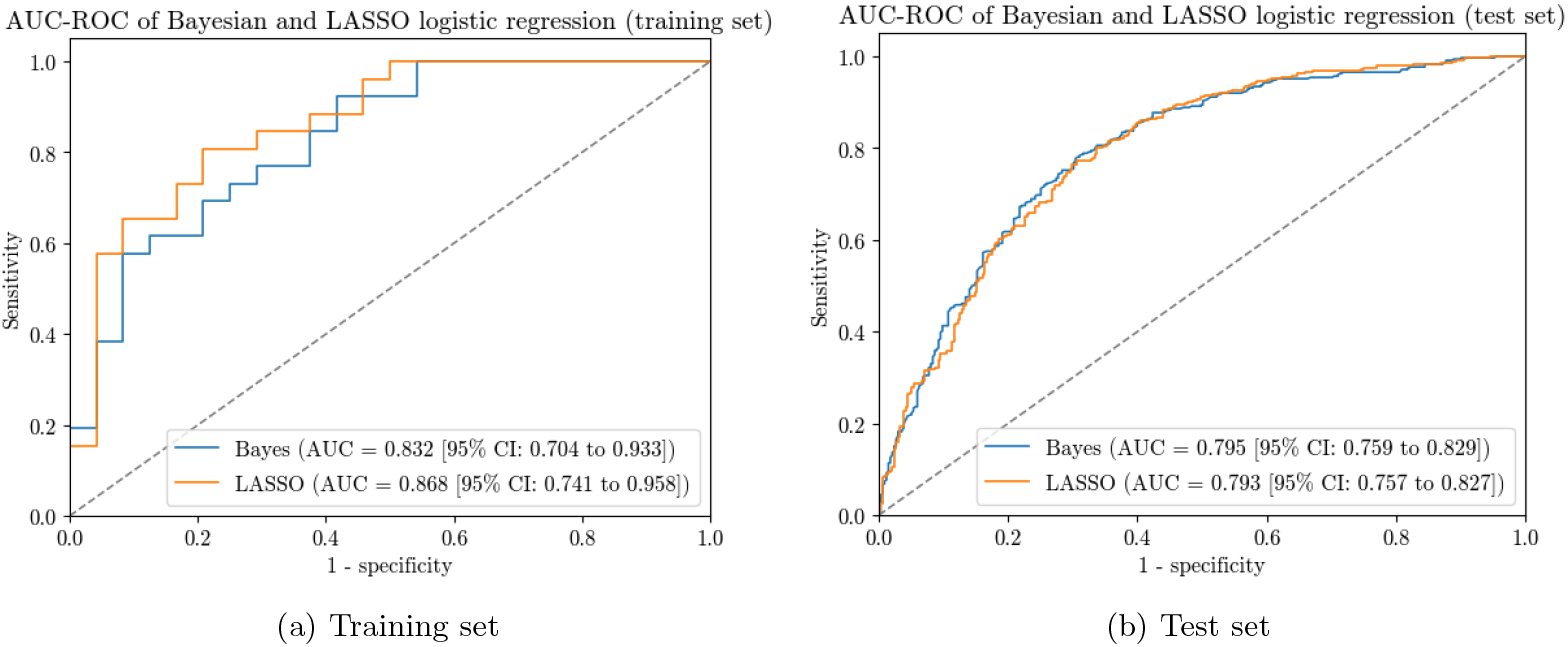
AUC-ROC plot of the Bayesian and Lasso logistic models on the (a) training set, (b) test set

In comparison, the Lasso logistic regression model achieved an AUROC score of 0.868 (95% CI: 0.741 to 0.958) on the training set and 0.793 (95% CI: 0.757 to 0.827) on the test set (Figure 1). The accuracy score for the Lasso model was 0.781 (95% CI: 0.660 to 0.900) on the training set and 0.728 (95% CI: 0.694 to 0.762) on the test set.

The Precision, Recall and F1-Scores of both models are summarized in Table 4.

**Table 4:** Precision, Recall, and F1-Scores of the Bayesian and Logistic regression models.

| Model | Classification | Precision (train, test) | Recall (train, test) | F1-Score (train, test) |
| --- | --- | --- | --- | --- |
| Bayes | Benign | 0.82, 0.81 | 0.58, 0.56 | 0.68, 0.66 |
|  | Malignant | 0.70, 0.68 | 0.88, 0.88 | 0.78, 0.76 |
|  | Macro average | 0.76, 0.74 | 0.73, 0.72 | 0.73, 0.71 |
| Lasso | Benign | 0.76, 0.74 | 0.79, 0.68 | 0.78, 0.71 |
|  | Malignant | 0.80, 0.72 | 0.77, 0.77 | 0.78, 0.74 |
|  | Macro average | 0.78, 0.73 | 0.78, 0.73 | 0.78, 0.73 |

**Abbreviations:** Bayes=Bayesian Logistic Regression; Lasso=Lasso Logistic Regression

## Discussion

This study demonstrates that SVSS, in conjunction with bayesian logistic regression, is a viable approach to radiomic-based small lung nodule classification. Generally, the performance of this approach on the that from the baseline Lasso logistic regression model, but the performance on the test set is similar (Figure 1 and Table 4).

The Bayesian approach excels in its ability to integrate prior information into the model via priors. For instance, the small *p* assigned to the Bernoulli prior in the SVSS model suggests that most features are irrelevant and should be excluded. The mildly informative priors used in this study reflect the uncertainty in feature selection during SVSS and the strength of coefficients in Bayesian logistic regression. In the SVSS model, reducing *τ* below 1 led to divergences during sampling, which is expected given the small sample size used for training, necessitating the specification of informative priors [7]. Although previous research [2] provided prior knowledge on promising candidate features, equal weight was assigned to all features to investigate whether SVSS would select different ones.

Indeed, out of the three features selected by SVSS, only Annulus GLCM Entrop LLL was common with the set of features selected by Hunter et al. [2]. This feature was consistently identified across all experiments and has generally been shown to increase malignancy prediction. Thus, future analyses using the bayesian approach should place greater emphasis on this feature to express confidence in it’s effect.

Another benefit of the Bayesian approach is it’s ability to quantify uncertainty in parameter estimates. Unlike traditional frequentist approaches, such as Lasso logistic regression, which typically provide point estimates and confidence intervals, Bayesian methods generate posterior distributions for each parameter. These parameter estimates represent the range of plausible values for the parameters, given prior information combined with the likelihood of observed data. This capability can offer valuable insights in the context of medical research, where decisions often need to be made under conditions of uncertainty. The HDI of the coefficients estimated from the bayesian logistic regression model was relatively large (Table 2). As more knowledge is acquired and increasingly informative priors are utilized for prediction, the coefficient estimates will improve.

The utility of Bayesian methods is particularly pronounced in settings with limited data. Traditional statistical methods often face challenges with small sample sizes, resulting in unstable and unreliable estimates. For instance, the univariate logistic regression models used in the initial layer of feature selection encounter convergence issues when the training sample size drops below 50. In contrast, Bayesian approaches effectively address this issue by integrating prior information, which stabilizes the estimates and improves model performance [7]. This characteristic makes Bayesian methods a powerful tool for radiomic analysis, where obtaining large datasets can be challenging due to the rarity of certain conditions or the high cost of imaging. Consequently, the Bayesian approach provides reliable and interpretable results in medical image classification tasks, such as the classification of radiomic-based small lung nodules.

Despite the advantages of incorporating prior information and improved interpretability, SVSS and Bayesian logistic regression can be computationally expensive, and scaling these methods to large datasets may present challenges. Furthermore, these methods are sensitive to the choice of priors, and poorly chosen priors can lead to biased or misleading conclusions. Therefore, careful consideration and tuning of priors are essential to ensure robust and accurate model performance. Additionally, leveraging advanced computational techniques, such as parallel processing and efficient sampling algorithms [1], can help mitigate the computational challenges and enhance scalability.

## Conclusion

This study aimed to explore the utility of Bayesian methods, specifically Stochastic Variable Selection and Shrinkage (SVSS) and Bayesian logistic regression, in radiomics-based classification of small lung nodules with limited training data. Although these methods could not be evaluated on a sample size smaller than 50 due to convergence issues in the univariate logistic regressions during the initial layer of feature selection, the Bayesian approach successfully matched the performance of frequentist Lasso logistic regression on the test set. This demonstrates its viability as an alternative approach.

Annulus_GLCM_Entrop_LLL was identified as a feature that consistently exerts a positive influence on small lung nodule malignancy prediction across various models. This finding enhances our confidence in the effect of this feature, suggesting that future Bayesian analyses can incorporate this information to achieve greater reliability in feature selection and coefficient estimates.

In conclusion, this study highlights the potential of Bayesian methods to address the challenges of limited data in medical image analysis. By leveraging prior information, these methods can stabilize estimates and improve model performance, offering a robust alternative to traditional statistical approaches. Future research should focus on further refining these Bayesian techniques and exploring their application in other high-dimensional radiomic datasets to enhance their utility and effectiveness in clinical decision-making.

## Data Availability

The radiomics data utilized in this study is publicly accessible in the Mendeley database under the accession code 10.17632/rxn95mp24d.1.
Additionally, the python scripts for model development are available at:
https://github.com/isaaclhk/Projects/tree/main/lung_nodule_SVSS

https://data.mendeley.com/datasets/rxn95mp24d/1

https://github.com/isaaclhk/Projects/tree/main/lung_nodule_SVSS

## Supplementary Materials

**Table S1:** SVSS diagnostic statistics.

| Feature | mcse_mean | mcse_sd | ess_bulk | ess_tail | $\hat{r}$ |
| --- | --- | --- | --- | --- | --- |
| Annulus_GLCM_Entrop_LLL_25HUGl | 0.006 | 0.007 | 3377.0 | 3377.0 | 1.0 |
| Annulus_GLRLM_SRLGLE_LLL_25HUGl | 0.005 | 0.007 | 3670.0 | 3670.0 | 1.0 |
| Annulus_GLRLM_LGLRE_LLL_25HUGl | 0.005 | 0.007 | 3565.0 | 3565.0 | 1.0 |

**Abbreviations:** mcse=Monte Carlo Standard Error; ess=Effective Sample Size

**Table S2:** Bayesian logistic regression diagnostic statistics.

| Coefficient | mcse_mean | mcse_sd | ess_bulk | ess_tail | $\hat{r}$ |
| --- | --- | --- | --- | --- | --- |
| intercept | 0.003 | 0.003 | 14770.0 | 11339.0 | 1.0 |
| beta[0] | 0.005 | 0.004 | 12576.0 | 12542.0 | 1.0 |
| beta[1] | 0.008 | 0.006 | 10920.0 | 11858.0 | 1.0 |
| beta[2] | 0.007 | 0.006 | 11710.0 | 11342.0 | 1.0 |

**Abbreviations:** mcse=Monte Carlo Standard Error; ess=Effective Sample Size

**Table S3:** Comparison of Bayesian Logistic Regression Models with varying prior parameters *τ*.

| $\tau$ | elpd_loo | p_loo | elpd_diff | Weight | se | dse |
| --- | --- | --- | --- | --- | --- | --- |
| 1 | -26.894 | 2.043 | 0.00 | 0.675 | 3.400 | 0.00 |
| 10 | -27.173 | 0.834 | 0.280 | 0.325 | 2.141 | 1.361 |
| 0.1 | -27.775 | 2.977 | 0.881 | 0.000 | 3.870 | 0.511 |
| 0.01 | -27.935 | 3.149 | 1.041 | 0.000 | 3.933 | 0.578 |

**Abbreviations:** elpd\_loo=Expected Log Pointwise Predictive Density for LOO-CV; p\_loo=Effective Number of Parameters; elpd\_diff=Difference in elpd\_loo; se=Standard Error; dse=Difference in Standard Error

**Figure S1:**
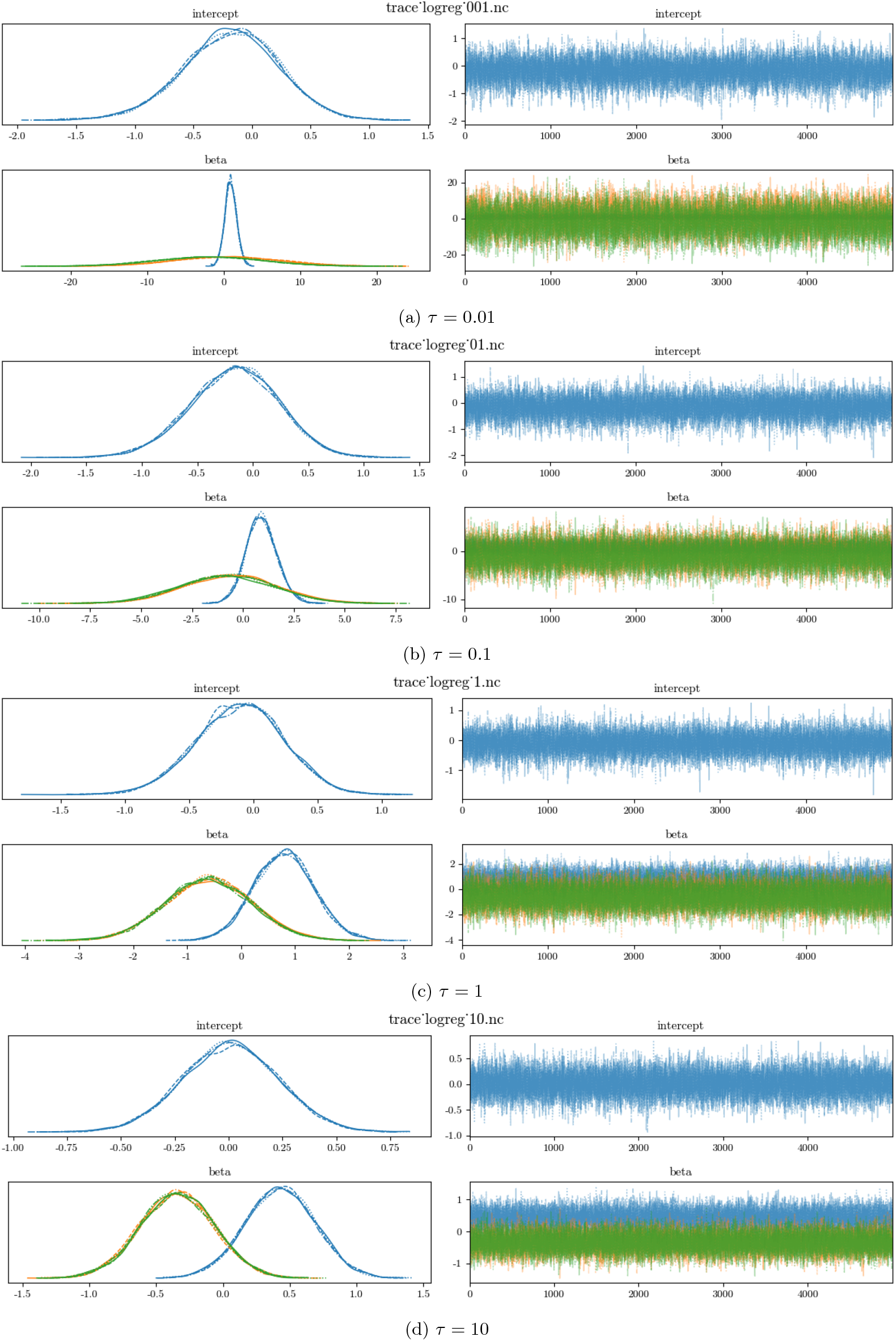
Bayesian Logistic Regression trace plots with *τ* =0.01 (a), 0.1 (b), 1 (c) and 10 (d)

## Notes

### Competing Interest Statement

The authors have declared no competing interest.

### Funding Statement

This study did not receive any funding

### Author Declarations

The dataset used in this study is available at the Mendeley database under the accession code 10.17632/rxn95mp24d.1. https://data.mendeley.com/datasets/rxn95mp24d/1

### Summary of Updates

The abstract was updated from an unstructured format to a structured format.

